# Modeling COVID-19 care capacity in a major health system

**DOI:** 10.1101/2021.11.18.21266407

**Authors:** Margret Erlendsdottir, Soheil Eshghi, Forrest Crawford

## Abstract

Hospital resources, especially critical care beds and ventilators, have been strained by additional demand throughout the COVID-19 pandemic. Rationing of scarce critical care resources may occur when available resource limits are exceeded. However, the dynamic nature of the COVID-19 pandemic and variability in projections of the future burden of COVID-19 infection pose challenges for optimizing resource allocation to critical care units in hospitals. Connecticut experienced a spike in the number of COVID-19 cases between March and June 2020. Uncertainty about future incidence made it difficult to predict the magnitude and duration of the increased COVID-19 burden on the healthcare system. In this paper, we describe a model of COVID-19 hospital capacity and occupancy that generates estimates of the resources necessary to accommodate COVID-19 patients under infection scenarios of varying severity. We present the model structure and dynamics, procedure for parameter estimation, and publicly available web application where we implemented the tool. We then describe calibration using data from over 3,000 COVID-19 patients seen at the Yale-New Haven Health System between March and July 2020. We conclude with recommendations for modeling tools to inform decision-making using incomplete information during future crises.

## 1 Introduction

The novel severe acute respiratory syndrome coronavirus (SARS COV-2), which causes coronavirus disease 2019 (COVID-19), emerged in 2019 in Hubei province in China. People infected with COVID-19 are at high risk for severe respiratory disease and serious complications [1]. Management of respiratory failure and acute respiratory distress syndrome (ARDS) often requires mechanical ventilation managed in an intensive care unit (ICU). In 2020 and the spring of 2021, the surge of COVID-19 patients in the United States revealed that critical care resources available in some health systems were insufficient to address the COVID-19 outbreak. Localized bed shortages occurred in parts of New York City [2], spurring hospital systems [3] and governors [4, 5] to order immediate bed capacity expansion. Failure to meet the critical care needs of COVID-19 patients has dire consequences; in Italy, reports of rationing of ventilators based on age cutoffs emerged from overwhelmed hospitals and ICUs [6, 7]. Furthermore, the spread of COVID-19 has affected the availability of staff [8].

The development and administration of vaccines such as Pfizer BNT162b2 and Modern mRNA-1273 in high-income nations like the United States, Israel, and the United Kingdom has slowed rates of infection with SARS-Cov-2, but critical care capacity continues to be overwhelmed by the unvaccinated population and the development of aggressive variants. As of fall 2021, one in four ICUs in the southern United States are above 95% capacity, with COVID19 patients accounting for approximately half of all ICU patients [9]. These high hospitalization rates are driven by evolving variants of COVID-19, such as the Delta variant, which the CDC has deemed more contagious than previous variants, more likely to cause severe illness, and capable of causing breakthrough infections in vaccinated people [10].

Tools for managing hospital capacity during surges in COVID-19 cases will be required as long as vaccination rates remain low in some countries and new variants continue to arise. Rapid practice guidelines recommend the use of mathematical modeling to guide surge capacity planning in hospital systems which expect to encounter potential shortages in critical care resources [11]. Guidelines state that the models should “be pragmatic and focus on the only relevant question for surge capacity: how many patients will need hospital and ICU resources on a given day?” More specifically, the models should provide early predictions, insight regarding both best and worst case scenarios, and the local rate of spread of infection and rate of hospitalization. Many modeling tools were created at the beginning of the COVID-19 pandemic to assist with predictions of incident COVID-19 cases and hospitalizations [12–25]. However, a comparison of four prominent models by Chin et al. [26] found that “for accuracy of prediction, all models fared very poorly.” These tools used population-level epidemic projections as inputs to their model of hospital occupancy, which may have contributed to compounding errors in forecasting hospital bed occupancy due to uncertainties in the early epidemiological models of COVID-19. The authors concluded that “trustworthy models require trustworthy input data” and that the models “need to be subjected to pre-specified real time performance tests.”

In this paper, we present a model of COVID-19 hospital occupancy that uses data from health systems to generate its predictions. The model is independent of the uncertainty in population-level epidemiological predictions of infection and can flexibly accommodate local variations important to decision-makers, such as significant differences in patient demographic distributions and hospital protocols. The model provides predictions of floor and ICU occupancy and mortality for infection scenarios specified by the hospital administrator or decision-maker. These infection scenarios can be informed both by observed presentations of COVID-19 patients to the health care system and epidemiological predictions of infection. The model can predict the effects of planned modifications to hospital capacity, and projections can be tailored to the dynamics of a specific hospital system using several parameters estimable from electronic health record data. We introduce the model structure and describe both the model dynamics and the calibration procedure for the model parameters. The model was calibrated using observed patient trajectories from the 3000-bed Yale New-Haven Hospital system, collected and processed during the surge in COVID-19 cases in Connecticut between March and July 2020. We validate the model dynamics using the observed hospital census during this time period. We conclude with recommendations to guide scientists developing model-based recommendations for managing hospital capacity during the COVID-19 pandemic.

## 2 Methods

### 2.1 Model structure

The goal of the model is to allow hospitals to generate projections of their occupancy and expected clinical outcomes. In this model, we describe the flow of COVID+ patients through a hospital system using a system of ordinary differential equations. Figure 1 shows the possible patient trajectories in the hospital system. By COVID+ patients, we refer to patients who have tested positive for COVID-19 prior to admission, those that test positive for COVID-19 within 14 days of admission, and transfers from other hospitals. We model transitions between eight different compartments: 1) presentations to the health system or emergency department, where triage occurs *P*, 2) floor beds *F*, 3) ICU beds *C*, 4) *MS*, corresponding to a state post-discharge from the emergency department

**Figure 1:**
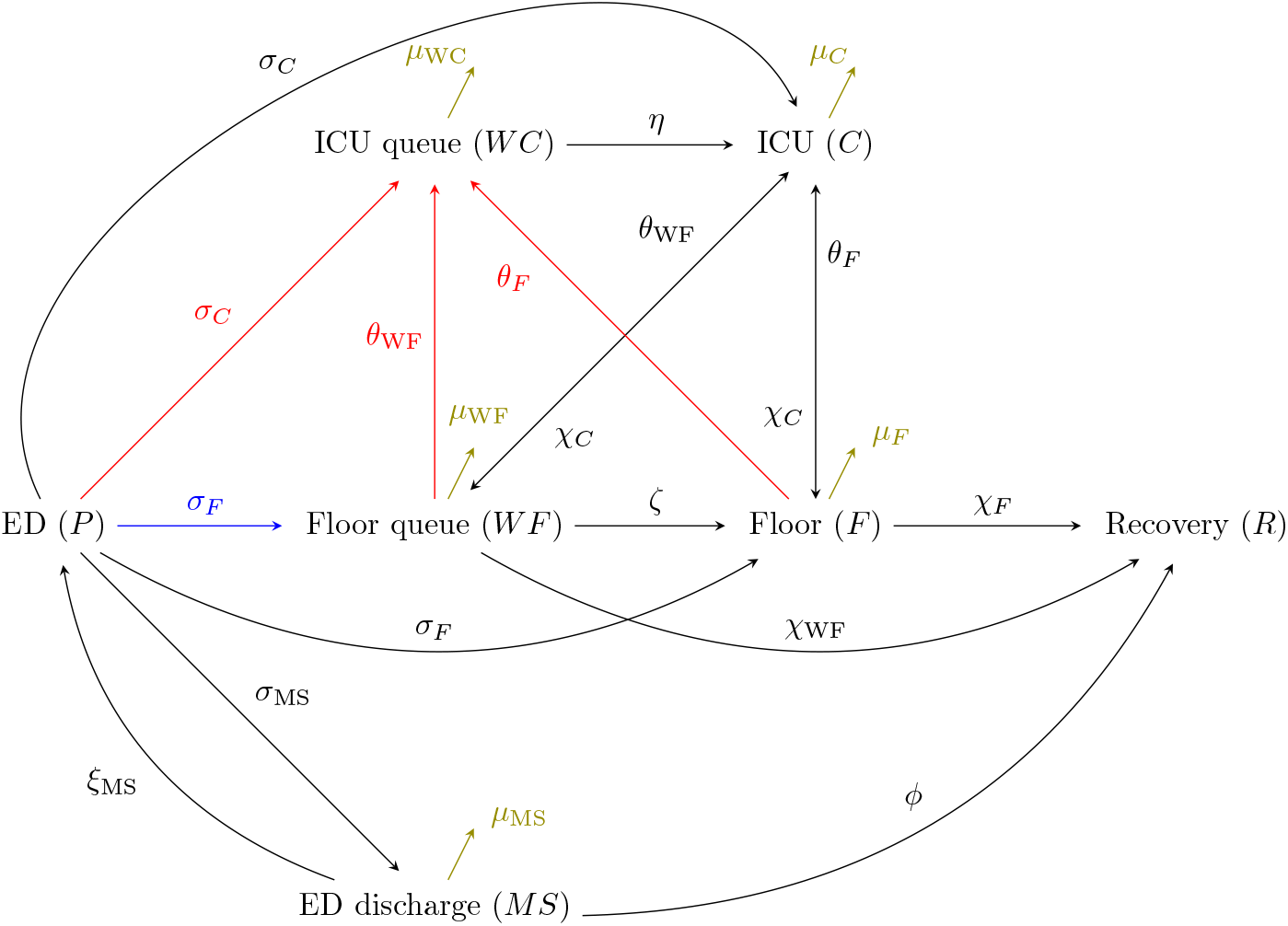
Model structure and parameters. Simplified dynamics are presented here for the case of one age-group for clarity. Patients present to the hospital system via the ED, where they are triaged and either discharged or admitted to the hospital. Patients who are admitted may go to the floor or directly to the ICU. The model captures patient flow from the floor to the ICU and back, as well as discharge dynamics from both the ED and the floor to recovery. Rate parameters which capture the speed at which patients transition between compartments are included here and described in Table 1. Arrows in red and blue represent patient-flow over-flow dynamics in the ICU and floor, respectively. Olive-colored arrows represent death rates from each compartment.

(ED) for those patients with mild symptoms, 5) *R*, corresponding to recovery post-discharge for patients admitted to the hospital, 6) *WF*, the queue for floor beds which would develop if floor beds are not available, 7) *WC*, the queue for ICU beds which would develop if ICU beds are not available, and 8) death.

The rate parameters associated with each step of the model are shown in Table 1. Governing equations are described fully in the Supplement.

**Table 1:**
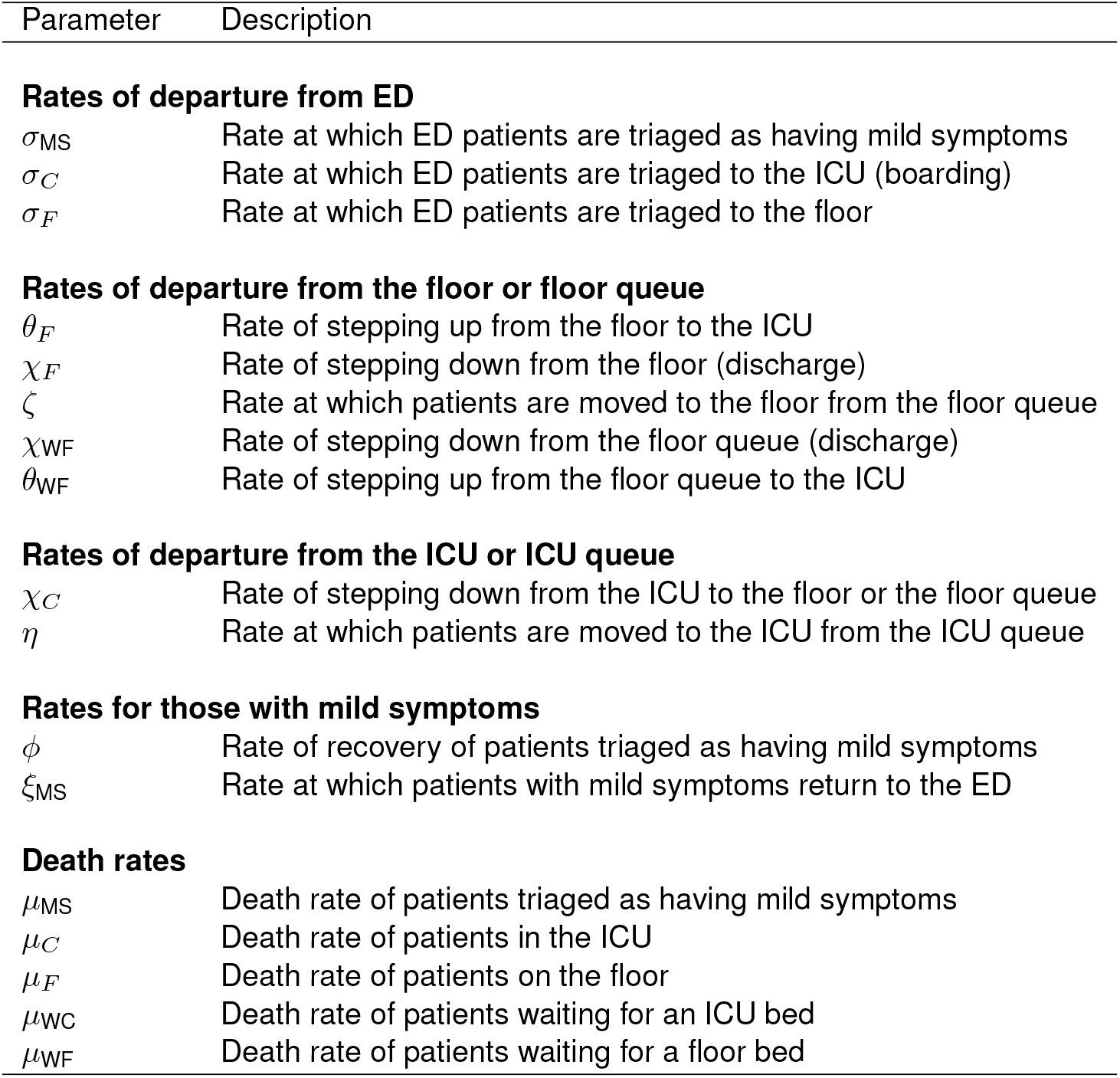
Rate parameters included in the model of hospital capacity Parameter Description

### 2.2 Specifying dynamics and capacity scenarios

The following features of the model are modifiable by the user in the web application in which the model is implemented.

The user begins by specifying an infection scenario: the number of COVID+ patients that present to a health system per day during a specified time horizon, between 2 and 60 days. They also select the number of COVID+ presentations at day zero of the projection and the expected change in the number of COVID+ presentations during the time of the simulation. For example, the user could choose an initial number of presentations of 50 patients, “exponential” change in patients, a doubling time of 14 days, and a time horizon of 14 days. This scenario corresponds to an exponential increase in the number of COVID+ presentations per day, such that 100 COVID+ patients present to the health system on day 14. Choices for the type of increase were: exponential, linear, saturated, and flat (no increase).

We allow the user to specify both the baseline number of available beds in the ICU and on the floor and a possible policy response, an increase their number of floor and ICU beds dedicated to COVID+ patients. The user may specify a one-time linear ramping of capacity.

The user may also modify several parameters which reflect the patient population in a specific catchment area, allowing the user to tailor the model to their particular needs. These parameters include the age distribution of admitted COVID+ patients, the average length of stay of COVID+ patients in the ICU and on the floor, and the probability of death of COVID+ patients in the ICU and on the floor. Default values in the web application are based on the YNHHS patient population.

Key outputs of the model which inform decision-marking regarding resource allocation are: the number of days to overflow, extra beds needed for COVID+ patients, number of deaths in each department, and predicted case-fatality rate. The ability to predict overflow could lead to dedication of non-COVID resources to COVID+ patients or the acquisition of additional space, for example. It is also clinically important, especially as the outcomes of patients that need ICU care are significantly worse if there are no available beds (a surge scenario). The predicted number of deaths among COVID+ patients in each department helps decision-makers understand the possible consequences of allocating beds and resources in different ways.

### 2.3 Model calibration

#### 2.3.1 Data sources

We used data from the Yale-New Haven Hospital System (YNHHS) collected between March 2020 and July 2020 to calibrate the model. YNHHS consists of five hospitals: Yale-New Haven Hospital (1,608 beds), Bridgeport Hospital (719 beds), Greenwich Hospital (304 beds), Lawrence and Memorial Hospital (260 beds), and Westerly Hospital (81 beds). For parameters which could not be estimated using available YNHHS data, we used population-level estimates from the Center for Disease Control’s Morbidity Mortality Weekly Report (CDC MWWR) [27–30]. This study received approval from the Institutional Review Board of Yale University’s Human Research Protection Program (IRB ID: 2000028666). We used three YNHHS data sources to calibrate the model: individual-level records for patients who had tested positive for COVID-19 in the YNHHS Emergency Departments (ED), individual-level records for patients who had been admitted to the hospital, and hospital-level summaries of capacity. Using these data, we reconstructed patient trajectories through the YNHHS hospitals. Figure 2A shows the total census of hospitalized COVID+ patients in YNHHS, as well as the census on the floor and ICU specifically. Figures 2B-E show the survival probabilities of death and departure from the floor and ICU, without accounting for competing hazards.

**Figure 2:**
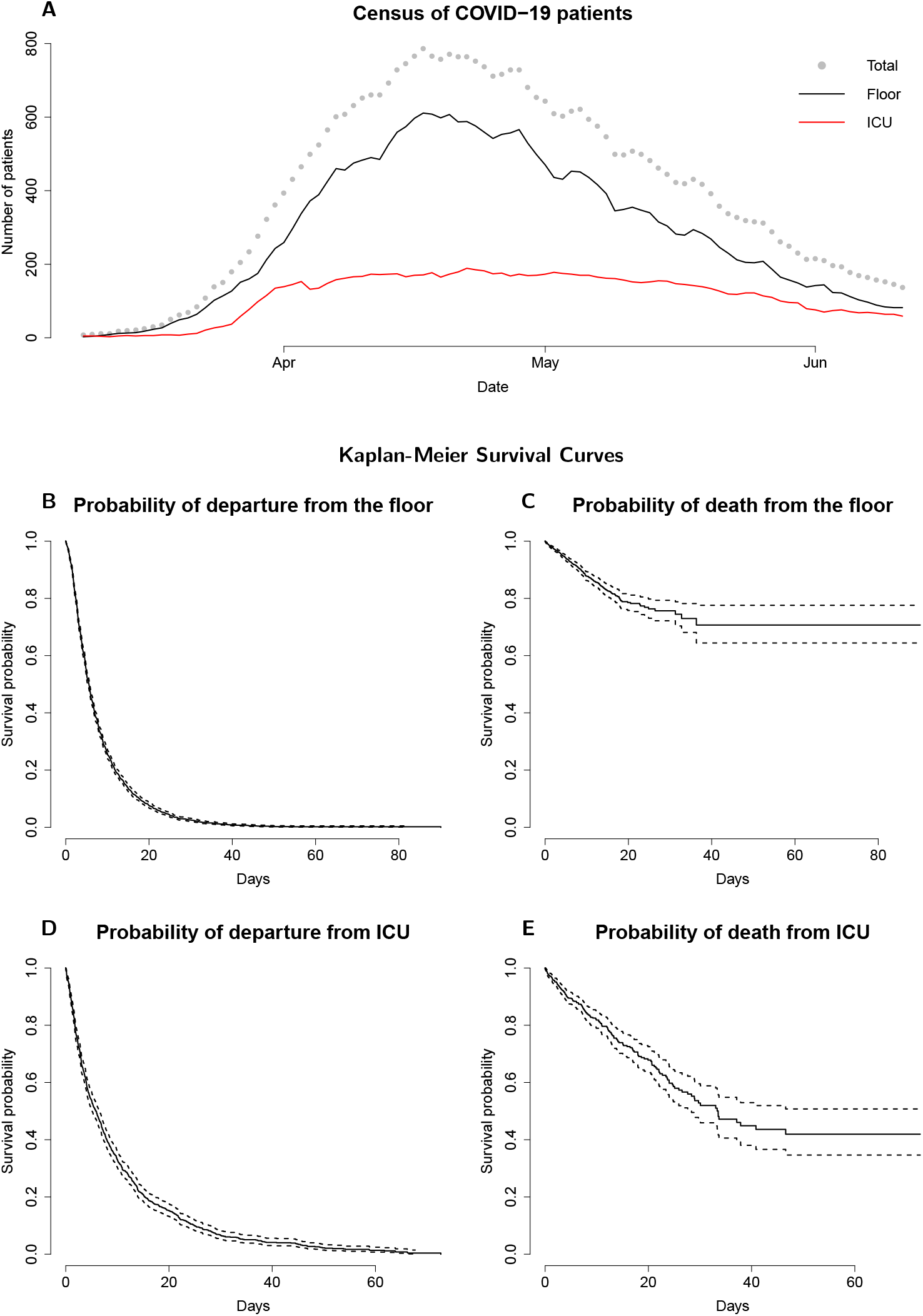
A: The census of hospitalized patients throughout YNHHS. The total census is shown with gray points, the census on the floor is shown in black, and the census in the ICU is shown in red. B and D: Kaplan-Meier curves describing the probability of remaining on the floor or ICU given time since arrival to the floor or ICU respectively. Patients are considered to be right-censored if they are still on the floor or in the ICU at the end of the observation period. Departure includes discharge, transfer to another department, and death. C and E: Kaplan-Meier curves describing the probability of survival, and not death, on the floor and in the ICU given time since arrival to the floor or ICU respectively. Patients are considered to be right-censored if they are transferred to another department, are discharged, or remain in the floor or in the ICU at the end of the observation period.

#### 2.3.2 Procedure for calibration of parameters

We used survival analysis with competing risks to estimate the following parameters governing rates of transition between hospital departments using the patient records available in the YNHHS dataset. We estimated three parameters describing rates of departure from the ED: from the ED to discharge (*σ*_*MS*_), the ED to admission to the floor (*σ*_*F*_), and the ED to admission to the ICU (*σ*_*C*_). We used inpatient data to estimate the rate of transition from the floor to the ICU (*θ*_*F*_), rate of discharge from the floor (*χ*_*F*_), rate of transition from the ICU to the floor (*χ*_*C*_), death while on the floor (*µ*_*F*_) and death while in the ICU (*µ*_*C*_). We performed a primary analysis in which time to all competing events were assumed to follow a gamma distribution. We estimated parameters in three age groups (0-17 years, 18-64 years, 65+ years). We used bootstrapping with 1,500 samples to generate estimates of the variance of these parameters. Additional statistical details of the procedure are described in the Supplement, as well as two secondary analysis two secondary analyses with simpler parameterizations to assess the performance of the estimated model parameters under different distributional assumptions.

### 2.4 Parameter estimates

The observed time-series of patients entering each department included both COVID-19 patients admitted directly to each department and direct transfers from other hospitals. Figure 2A shows the total census of COVID-19 patients admitted to all YNHHS hospitals during the observation period. Parameter estimation used all observed YNHHS patient trajectories, and model fit was evaluated using the the largest hospital, YNHH, where decision-making regarding resource allocation was most critical. In total, 2,275 COVID-19 patients who met criteria were admitted or transferred to YNHH during the observation period. The estimated capacity of YNHH for COVID-19 patients was 180 beds in the ICU and 578 beds on the floor. YNHH neither reached capacity nor ran out of ventilators during the surge in COVID-19 cases.

We used individual-level patient trajectories and time-stamped transitions between departments to calculate the time spent by each patient in each department, as well as the destination of each patient following departure. The estimated rates, assuming gamma-distributed time to event, are listed in Table 2. Using these estimated rates, we computed transition probabilities between the ED, floor, and ICU, and lengths of stay in each department; we include these transformed values in Table 3. Average length of stay (LOS) on the floor was 10 days (95% CI: 9.0, 11) for those over the age of 64 and 7.6 days (95% CI: 7.1, 8.1) for those between 18-64 years. LOS in the ICU was longest on average, 14 days (95% CI: 12, 116)) for adults between 19-64 years, 11 days (95% CI: 9.7, 13) for adults over 65 years, and 8.5 days (95% CI: 1.5-15) for children under the age of 18.

**Table 2:** Model parameters: Included in this table are both estimated rates of transition with 95% confidence intervals, rates taken from external sources [27–30] and rates which we set after being unable to determine then either from data or the literature. The estimated rates are based on the assumption that the time to each competing risk follow a two-parameter gamma distribution, where the product of the two parameters yield the estimated mean of the distribution. These parameters were estimated separately for each age group. Rates labeled with (*) were taken from CDC MMWR [27].

|  | 0-18 years | 18-64 years | 65+ years |
| --- | --- | --- | --- |
| **Estimated parameters** |  |  |  |
| $\sigma_{MS}$ | 1.1 (0, 4.1) | 0.081 (0, 1.9) | 0.55 (0.43, 0.68) |
| $\sigma_C$ | 0.017 (0, 0.32) | 0.075 (0, 0.67) | 1.1 (0.87, 1.2) |
| $\sigma_F$ | 0.37 (0, 2.3) | 0.035 (0, 2.4) | 3.4 (3.2, 3.6) |
| $\chi_C$ | 0.12 (0.011, 0.27) | 0.061 (0.05, 0.072) | 0.064 (0.052, 0.076) |
| $\chi_F$ | 0.28 (0.17, 0.4) | 0.11 (0.11, 0.12) | 0.071 (0.068, 0.075) |
| $\chi_{WF}$ | 0.28 (0.17, 0.4) | 0.11 (0.11, 0.12) | 0.071 (0.068, 0.075) |
| $\theta_F$ | 0.0095 (0, 0.041) | 0.014 (0.011, 0.017) | 0.013 (0.011, 0.016) |
| $\theta_{WF}$ | 0.0095 (0, 0.041) | 0.014 (0.011, 0.017) | 0.013 (0.011, 0.016) |
| $\zeta$ | 2.7 (1.5, 4) | 1.2 (1.1, 1.3) | 0.87 (0.79, 0.95) |
| $\xi_{MS}$ | 0.012 (0.012, 0.012) | 0.0054 (0.0054, 0.0054) | 0.0039 (0.0039, 0.0039) |
| $\mu_{MS}$ | 0.00066 (0.00066, 0.00066) | 0.00066 (0.00066, 0.00066) | 0.00066 (0.00066, 0.00066) |
| $\mu_C$ | 0.0035 (0.0023, 0.0048) | 0.0083 (0.0038, 0.013) | 0.024 (0.018, 0.03) |
| $\mu_F$ | 0.002 (0.0018, 0.0022) | 0.00033 (0, 0.001) | 0.012 (0.0092, 0.015) |
| $\mu_{WF}$ | 0.002 (0.0018, 0.0022) | 0.00033 (0, 0.001) | 0.012 (0.0092, 0.015) |
| **Fixed parameters** |  |  |  |
| $\phi$ | 0.088* | 0.094* | 0.095* |
| $\mu_{WC}$ | 4 | 4 | 4 |
| $\eta$ | 36 | 36 | 36 |

**Table 3:** Probabilities of transition between the ED, Floor, and ICU, and lengths of stay: Included in this table are both estimated probabilities of transition with 95% confidence intervals, and lengths of stay in each hospital department. As with the estimated rates in Table 2, the estimated probabilities and lengths of stay are based on the assumption that the time to each competing risk follow a two-parameter gamma distribution, where the product of the two parameters yield the estimated mean of the distribution. These parameters were estimated separately for each age group.

|  | 0-18 yrs | 19-64 yrs | 65+ yrs |
| --- | --- | --- | --- |
| Age distribution in ED, % | 0.02 (0.02, 0.02) | 0.58 (0.58, 0.58) | 0.4 (0.4, 0.4) |
| % discharged from ED | 0.73 (0.56, 0.9) | 0.54 (0.33, 0.75) | 0.11 (0.09, 0.13) |
| % admitted from ED to floor | 0.25 (0.1, 0.4) | 0.26 (-0.053, 0.57) | 0.68 (0.65, 0.71) |
| % admitted from ED to ICU | 0.037 (-0.02, 0.052) | 0.2 (0.095, 0.31) | 0.21 (0.19, 0.24) |
| % death on the floor | 0.024 (0.015, 0.033) | 0.0052 (-0.00013, 0.011) | 0.13 (0.1, 0.15) |
| % death in the ICU | 0.01 (0.0069, 0.013) | 0.13 (0.069, 0.18) | 0.27 (0.22, 0.32) |
| % step up from floor to ICU | 0.52 (0.22, 0.81) | 0.79 (0.73, 0.85) | 0.65 (0.59, 0.7) |
| % step down to the floor | 0.042 (-0.034, 0.12) | 0.11 (0.089, 0.13) | 0.14 (0.12, 0.16) |
| Triage time in ED (days) | 0.81 (-0.17, 1.8) | 4 (-1.2, 9.3) | 0.2 (0.18, 0.21) |
| Average LOS on floor (days) | 3.3 (2, 4.6) | 7.6 (7.1, 8.1) | 10 (9.6, 11) |
| Average LOS in ICU (days) | 8.5 (1.5, 15) | 14 (12, 16) | 11 (9.7, 13) |

### 2.5 Model fit

Accurate predictions of occupancy were the most important output of the model, as a tool to help hospital administrations with surge planning. We evaluated the ability of the model to generate predictions of hospital occupancy at YNHH that matched the observed occupancy during a surge in COVID-19 patients at Yale-New Haven Hospital (YNHH), the largest of the five hospitals in YNHHS. These predictions were generated using the observed time series of patients admitted to each YNHH department between March 8 and June 12, 2020, and were compared to the observed occupancy in each department during this time. Parameters were estimated assuming gamma-distributed time to departure from a department. Figure 3 show the observed occupancy and occupancy predicted by the model according to this analysis.

**Figure 3:**
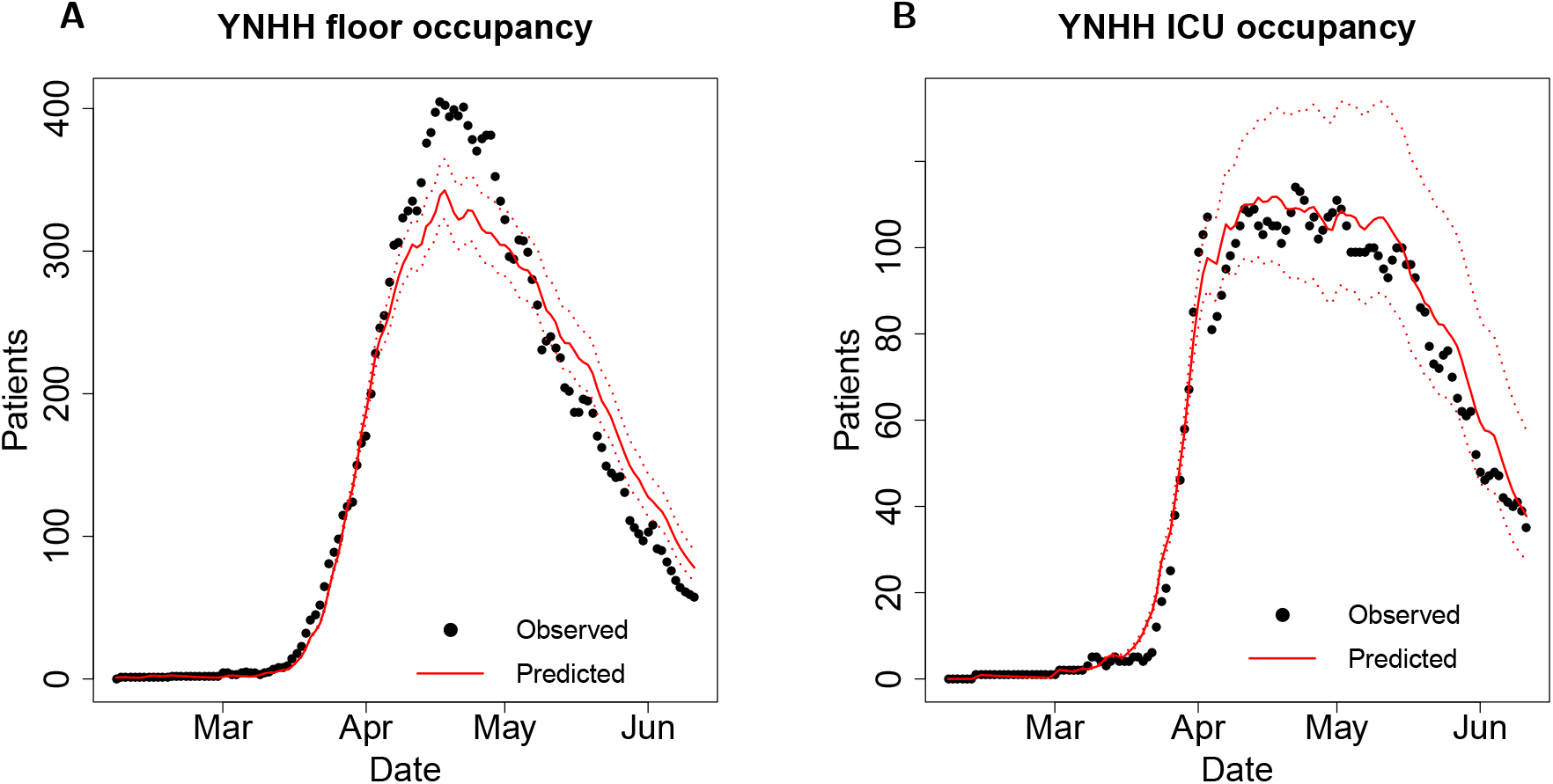
Inpatient predicted and observed COVID19 floor occupancy (A) and ICU occupancy (B). Parameters describing rates of transition between hospital departments were estimated assuming gamma-distributed time to event. The dotted line represents occupancy at YNHH. The solid red line represents model output based on parameters calculated using our fitting procedure and capacity estimates from YNHH. The dotted red lines represent estimated occupancy according to the bounds of 95% confidence intervals for each parameter.

The model accurately predicts occupancy in YNHH. Figure 3B shows that the ICU occupancy predicted by the model closely matches the increase in the observed YNHH ICU occupancy between March and April, the period of peak occupancy between April and May, and the decrease in occupancy between May and June. Similarly, Figure 3A demonstrates that the model also accurately predicts the increase in floor occupancy between March and April and the date of peak occupancy, April 18, 2020. The model predicts that floor occupancy on April 18 would be 343 patients (95% CI: 323, 365). The observed YNHH occupancy on April 18 was 405 patients. The model’s prediction for deaths among COVID-19 patients on the floor and in the ICU were also reasonable but are not shown for privacy reasons.

## 3 Discussion

In this paper, we have presented a model of hospital capacity which was utilized by YNHHS to inform decisionmaking regarding resources which would be necessary to handle the surge in COVID-19 patients at YNHHS. As requested by rapid response guidelines [11], the model predicts the number of patients requiring hospital resources under a variety of scenarios. We have described the process that we used to fit the model using data from YNHHS collected as the surge in COVID-19 patients was occurring. We have provided evidence that our estimation procedure provides a reasonable estimate of the true dynamics in a system by comparing the performance the model to the observed dynamics in YNHH during the surge in COVID-19 cases.

The model successfully predicted the most important quantity: occupancy in the ICU, which is the most scarce resource and most important for helping severely ill COVID-19 patients. However, this method may have several weaknesses. The model slightly underpredicted floor occupancy. This could be due to inaccurate assumptions used to estimate the model parameters. The model parameters were estimated for 3 age groups. The model fit could be improved in future versions by creating additional subgroups according to age and other risk factors for hospitalization and severe disease in COVID-19 patients. The model also assumes that rates of transition between departments remains constant over time. However, several factors, including changing hospital protocols for triage and treatment, may have resulted in fluctuations in the rates of transition over time. Such non-stationary behavior is challenging to capture and would not be captured by the model or parameter estimation procedure. In addition, the model is deterministic, and the estimates of variance in occupancy are based on uncertainty in parameter estimation rather than inherent stochasticity in the model. Improved estimates of variance might be achieved by making the model fully stochastic.

The urgency of the crisis caused by surging COVID-19 patients contributed substantially to the challenge of developing a useful model. We would like to conclude by providing a few recommendations for creating a model in a crisis. First, early collaboration with end-users of the product was essential. After creation of the model structure and early implementation of the web application, we met several times with administrators at YNHHS who were in charge of capacity planning. They provided feedback on the model and the web application, in addition to crucial perspective on the most urgent unmet needs which could be addressed by the model. Second, we recommend reducing the dependence of these models on unverifiable assumptions. Instead of constructing a population-level model which would predict hospitalizations without explicitly modeling dynamics within a hospital, almost all of the parameters used to fit this model are based on observable data from electronic health records. Despite using observable data to construct the model, we still had to consider the implications of the incomplete nature of the dataset. The model was sensitive to differences in parameters that were estimated using different procedures. The fidelity in the predictions of ICU occupancy was only achieved after we took into account right-censoring of patient trajectories, with sicker patients remaining in the hospital at the time of the analysis. Third, interactive implementations of any model results should be streamlined to involve the smallest possible number of parameters for ease of use, and the rest should be reasonable defaults. Despite the large number of parameters necessary to use the model, we included only a limited number for users of the web application to manipulate.

## Data Availability

Data utilized in the present study are not available because they are protected health information. Code produced in the study is available upon request.

https://forrestcrawford.shinyapps.io/covid19_icu/

## Acknowledgments

This work was supported by NIH grant NICHD DP2 HD091799-01. We would like to thank Amento Annette, Thomas Balcezak, Keisha Boykin, Matthew Comerford, Philip Corso, Rick D’Aquila, Gary Desir, Edward Kaplan, Brian Keane, Thomas Prem, Rema Seeram, and Paul Taheri at the Yale New Haven Hospital system for their support and feedback. We would also like to acknowledge the COVID-19 Statistics, Policy Modelling and Epidemiology Collective (C-SPEC), which included the following individuals: David Paltiel, Gregg Gonsalves, Jeffrey Eaton, Josh Salomon, Meagan Fitzpatrick, Nick Menzies, Reza Yaesoubi, Adam Beckman, Alyssa Bilinski, Anna York, Anne Williamson, Bianca Mulaney, Christian Testa, Emma Clarke-Deelder, Evan-MacKay, Hanna Ehrlich, Jinyi Zhu, John Giardina, Katie Rich, Kayoko Shioda, Lin Zhu, Elizabeth White, Luke Massa, Maile Phillips, Melanie Chitwood, Monica Farid, Nicole Swartwood, Nikhil Deshmukh, Raphael Sherak, Ruthie Birger, Samantha Burn, Stephanie Perniciaro, Suzan Iloglu, Thomas Thornhill, Tyler Copple, Yu-Han Kao, and Yuli-Lily Hsieh. The web application was written by Soheil Eshghi, Margret Erlendsdottir, Maile Thayer Phillips, Suzan Iloglu, Christian Testa and Forrest W. Crawford using the R shiny framework. We are especially grateful to Gregg Gonsalves, David Paltiel, Hanna Ehrlich, Raphael Sherak, Melanie Chitwood, Thomas Thornhill, Nicole Swartwood, and Stephanie Perniciaro for advice and comments.

## Conflicts of interest

We have no conflicts of interest to report.

### Data statement

All data are protected health information and are not available for sharing. Code is available on request.

## Notes

### Competing Interest Statement

FWC has received consulting fees from Revelar Biotherapeutics and Whitespace Ltd.

### Funding Statement

The work was funded by NICHD grant 1DP2HD091799-01.

### Author Declarations

The Yale University Institutional Review Board gave ethical approval for this work.

