## Supplement for "Modeling COVID-19 care capacity in a major health system"

#### 1 Model structure and governing equations

We model the following events in the health system.

- **Presentation to the health system:** COVID+ patients can present to their health system for asymptomatic COVID-19 screening, following a positive COVID-19 test, or following initial onset of symptoms. They may present either to outpatient clinics or to the ED of their local hospital. At these locations, *triage* occurs, such that patients are either discharged with mild symptoms, admitted to the floor, or admitted to the ICU.
- **Following admission to the floor:** Patients admitted to the floor can be discharged directly from the floor, require *stepping up* to an ICU due to a deterioration in their condition, or die on the floor. Patients may arrive on the floor from the ED or after leaving the ICU following an improvement in their condition.
- **Following admission to the ICU:** Patients in the ICU require frequent monitoring and intensive interventions. COVID+ patients are especially at risk of requiring critical care due to high rates of pneumonia and acute respiratory distress syndrome (ARDS) reported in COVID+ patients [1–6]. Following recovery, ICU patients *step down* to the floor for discharge. Patients may also die in the ICU.
- **Following discharge from the ED:** If a patient's condition is not severe, COVID+ patients are instructed to return home and to self-isolate. Most patients with mild disease recover during their isolation. Some patients return to the ED in worsened condition, and a small number may die after discharge.
- **If the floor has reached capacity:** If floor beds are not available, patients may have to wait in the ED or elsewhere until a bed becomes available. During this time, patients are often monitored or receive care. They may be discharged if they recover, or die.
- **If the ICU has reached capacity:** If ICU beds are not available, patients may have to wait on the floor, in the ED, or elsewhere until a bed becomes available. Because these patients are critically ill, the probability of death if no care if received is high.

We stratify incoming patients into age tiers, based on known differences in patient outcomes by age [3]. Dynamics for each age group are governed by the following system of ordinary differential equations. The dynamics for each age group are coupled by the constraint on total hospital floor beds $L$ and total ICU beds $M$. For age group $i$,

$$\frac{dP_i}{dt} = \xi_{MS_i} MS_i - (\sigma_{MS_i} + \sigma_{C_i} + \sigma_{F_i} + \mu_{P_i}) P_i$$

$$\frac{dMS_i}{dt} = \sigma_{MS_i} P_i - (\phi + \mu_{MS_i} + \xi_{MS_i}) MS_i$$

$$\frac{dWC_i}{dt} = (\sigma_{C_i} P_i + \theta_{F_i} F_i + \theta_{WF_i} WF_i) \left(1 - \frac{1}{1+e^{s(C-M)}}\right) - \mu_{WC_i} WC_i - \eta WC_i \left(\frac{1}{1+e^{s(C-M)}}\right)$$

$$\frac{dC_i}{dt} = (\sigma_{C_i} P_i + \theta_{F_i} F_i + \theta_{WF_i} WF_i + \eta WC_i) \left(\frac{1}{1+e^{s(C-M)}}\right) - (\mu_{C_i} + \chi_{C_i}) C_i$$

$$\frac{dWF_i}{dt} = (\sigma_{F_i} P_i + \chi_{C_i} C_i) \left(1 - \frac{1}{1+e^{s(F-L)}}\right) - \zeta WF_i \left(\frac{1}{1+e^{s(F-L)}}\right) - (\mu_{WF_i} + \theta_{WF_i}) WF_i$$

$$\frac{dF_i}{dt} = (\sigma_{F_i} P_i + \zeta WF_i + \chi_{C_i} C_i) \left(\frac{1}{1+e^{s(F-L)}}\right) - (\chi_{F_i} + \mu_{F_i} + \theta_{F_i}) F_i$$

$$\frac{dR_i}{dt} = \phi MS_i + \chi_{F_i} F_i$$

$$\frac{dD_i}{dt} = \mu_{MS_i} MS_i + \mu_{WC_i} WC_i + \mu_{C_i} C_i + \mu_{WF_i} WF_i + \mu_{F_i} F_i,$$

where $I_i(t)$ represents initial COVID+ presentations to the ED of age group $i$, $F = \sum_i F_i$ is the occupied floor capacity, $C = \sum_i C_i$ is the occupied ICU capacity, $L$ and $M$ are their respective available capacities, and $D$ is the state of death. The exponential sigmoidal terms in the above system of equations represent approximations to the ideal on-off switch, i.e., $f(x) = \mathbf{1}_{x \geq i} \approx (1 - \frac{1}{1+e^{s(x-i)}})$, with the parameter $s$ controlling the fidelity of the approximation, the higher, the better. The use of sigmoids speeds up the computation of the solution of the system of equations significantly, allowing the user to rapidly observe the effect of changes in inputs on the outputs of the model with minimal error. A graphical depiction of these dynamics is shown in Figure **??**.

#### 2 Parameter estimation

To estimate model rate parameters, we estimated the exit rate of patients from each compartment of the model, assuming exponentially distributed length of stay (LOS) in each compartment. We estimated exit rates for each competing hazard. For patients presenting to a YNHHS ED, we had access to ED arrival time, the primary chief complaint, age, ED departure time, admission status, admission department, the time of the positive COVID-19 laboratory test, and the date and time of previous presentation to the ED. For patients who were admitted, we had access to daily records including inpatient departments, location, age, and the dates at which they were moved between departments, admitted to the ICU, initiated on ventilation, discharged, or died.

We estimated the rates of each of these competing hazards by constructing a likelihood function in the following way. For each competing hazard $k$, $k = 1, .., K$, the time until event $k$ is $T_k$, and is exponentially distributed with rate $\lambda_k$. We assume independence between all $T_k$. We observe $T = \min_k T_k$, an indicator $\delta$ which takes value $k$ if the subject experienced event $k$, and a censoring indicator $C$ which takes value 1 if a subject remains in the hospital at the end of the study period and is 0 otherwise. We have $N$ total subjects in our dataset, and for each individual $i$, $i = 1, ..., N$, the observed data $O_i = (t_i, \delta_i, C_i)$.

We constructed a likelihood function to estimate the rate parameters $\lambda_k$ for each of $T_k \sim \exp(\lambda_k)$ or shape and scale parameters $\alpha_k$ and $\beta_k$ if $T_k \sim$ Gamma$(\alpha_k, \beta_k)$. Each set of distributional assumptions utilizes a different set of parameters, so we denote generally the vector of parameters used in a particular likelihood with $\Theta$ and the parameters describing the distribution of $T_k$ with $\theta_k$. We performed three analyses using different distributional assumptions for the density $f_k(t; \theta_k)$. The first analysis assumed that all $T_k$ were exponentially distributed. The second analysis assumed that all $T_k$ were exponentially distributed, except for those corresponding to discharge, which were gamma distributed. The third analysis assumed two-parameter gamma distributions for all $T_k$. As is typical in survival analysis, the density of time to event $T_k$ is $f_k(t;\theta_k)$, the survival function for event type $k$ is $S_k(t;\theta_k) = (1 - F_k(t;\theta_k))$, the hazard of event $k$ is $h_k(t;\theta_k) = f_k(t;\theta_k)/S_k(t;\theta_k)$, the overall survival function given independence between events is $S(t;\Theta) = \prod_{k=1}^{K}(1 - F_k(t;\theta_k))$. The contribution of an uncensored individual $i$ with an observed outcome is:

$$f_k(t_i;\theta_k) = h_k(t_i;\theta_k)\prod_{k=1}^{K} S_k(t_i;\theta_k) = f_k(t_i;\theta_k)\prod_{j\neq k}(1 - F_j(t_i;\theta_k))$$

The contribution of a censored individual is simply $\prod_{k=1}^{K}(1 - F_j(t_i;\theta_k))$.

Thus, the likelihood function for the observed data is:

$$L(t_1, ..., t_N, \Theta) = \prod_{i=1}^{N}\Big[\prod_{k=1}^{K} f_k(t_i;\theta_k)\prod_{j\neq k}(1 - F_j(t_i;\theta_k))\Big]^{\mathbb{I}(\delta_i = k)(1-C_i)}\prod_{k=1}^{K}(1 - F_j(t_i;\theta_k))^{C_i} \quad (1)$$

For all instances in which a gamma distribution was assumed, we computed the estimated mean of each $T_k$ using the maximum likelihood parameter estimates. These estimates, in addition to estimates obtained assuming exponential distributions, were together used to estimate the probability of transition out of each department and length of stay in the ICU and on the floor in the following away. We assumed all $T_k$ to be exponentially distributed random variables with rate $\lambda_k$, including those parameters estimated assuming gamma distribution in the log likelihood. If $X$ is the random variable which denotes the event which occurs, the probability of transition to a particular event $k$ is $\mathbb{P}(X = k) = \frac{\lambda_k}{\sum_k \lambda_k}$. The length of stay within a department is also exponentially distributed, with $\mathbb{E}[T]$ = $\mathbb{E}[\min_k T_k] = \frac{1}{\sum_k \lambda_k}$. For the purposes of parameter estimation, discharge from the ICU and step down from the ICU were considered to be separate competing hazards. The estimated probability of step down from the ICU used to parameterize the model is the sum of the probabilities of step down and discharge. Due to the relatively small number of people discharged directly from the ICU, we considered this to be a reasonable modification.

Several parameters could not be estimated from available YNHHS data. In most cases, these parameters were estimated using data published in the CDC MMWR [3–6]. The YNHHS data did not provide information regarding probabilities or times to full recovery among individuals after they were discharged. Therefore, rates of recovery ($\phi$), and death rates among individuals with mild symptoms ($\mu_{MS}$) were calculated in each age group from population-level proportions provided by CDC MMWR [3]. Because queues for floor beds and ICU beds did not occur at YNHHS hospitals at the time of model implementation, we were required to make several assumptions to generate the remaining parameters. We estimated that the rate of death, discharge, and step up to the ICU from the floor queue ($\mu_{WF}$, $\chi_{WF}$, and $\theta_{WF}$, respectively) would be the same as death and discharge rates from the floor, due to the fact that many hospitals are able to provide care to patients who are waiting for a floor bed before one becomes available. We set the average time of death in the ICU queue without access to critical care resources to be 6 hours. We set the probability of movement from the floor queue to an open floor bed and the ICU queue to an open ICU bed to be 0.9, reflecting a 90% chance that a patient would move from the queue to an open bed before discharge, death, or transition to another department. Rates of movement out of the queues were calculated accordingly.

#### Figures and tables for alternative distributional assumptions in parameter estimation procedure.

##### 1. Exponential model:

| | 0-18 years | 18-64 years | 65+ years |
|---|---|---|---|
| $\phi$ | 0.088 (0.088, 0.088) | 0.094 (0.094, 0.094) | 0.095 (0.095, 0.095) |
| $\sigma_{MS}$ | 1.4 (0, 5) | 2.4 (2.2, 2.6) | 0.43 (0.36, 0.49) |
| $\sigma_C$ | 0.072 (0, 0.33) | 0.4 (0.34, 0.47) | 0.48 (0.42, 0.55) |
| $\sigma_F$ | 0.48 (0, 1.8) | 2.1 (2, 2.3) | 3.1 (2.9, 3.2) |
| $\chi_C$ | 1.6 (1.6, 1.6) | 0.071 (0.06, 0.081) | 0.072 (0.061, 0.082) |
| $\chi_F$ | 1.6 (1.6, 1.6) | 0.11 (0.1, 0.12) | 0.065 (0.062, 0.068) |
| $\chi_{WF}$ | 1.6 (1.6, 1.6) | 0.11 (0.1, 0.12) | 0.065 (0.062, 0.068) |
| $\theta_F$ | 7 (7, 7) | 0.017 (0.014, 0.02) | 0.015 (0.014, 0.017) |
| $\theta_{WF}$ | 7 (7, 7) | 0.017 (0.014, 0.02) | 0.015 (0.014, 0.017) |
| $\eta$ | 36 | 36 | 36 |
| $\zeta$ | 78 (78, 78) | 1.2 (1.1, 1.3) | 0.86 (0.8, 0.92) |
| $\xi_{MS}$ | 0.012 (0.012, 0.012) | 0.0054 (0.0054, 0.0054) | 0.0039 (0.0039, 0.0039) |
| $\mu_{MS}$ | 0.00066 (0.00066, 0.00066) | 0.00066 (0.00066, 0.00066) | 0.00066 (0.00066, 0.00066) |
| $\mu_C$ | 2.7e-07 (0, 1.1e-06) | 0.0083 (0.0061, 0.011) | 0.03 (0.025, 0.035) |
| $\mu_F$ | 5.1e-07 (4.7e-07, 5.6e-07) | 0.0024 (0.0015, 0.0033) | 0.016 (0.014, 0.018) |
| $\mu_{WC}$ | 4 | 4 | 4 |
| $\mu_{WF}$ | 5.1e-07 (4.7e-07, 5.6e-07) | 0.0024 (0.0015, 0.0033) | 0.016 (0.014, 0.018) |

Table 1: Parameters assuming exponential distributions for each competing risk.

| | 0-18 yrs | 19-64 yrs | 65+ yrs |
|---|---|---|---|
| Age distribution in ED | 0.02 (0.02, 0.02) | 0.58 (0.58, 0.58) | 0.4 (0.4, 0.4) |
| % discharged from ED | 0.71 (0.62, 0.81) | 0.49 (0.47, 0.51) | 0.11 (0.092, 0.12) |
| % admitted from ED to floor | 0.25 (0.16, 0.34) | 0.43 (0.41, 0.45) | 0.77 (0.75, 0.79) |
| % admitted from ED to ICU | 0.037 (-0.0032, 0.078) | 0.081 (0.07, 0.092) | 0.12 (0.11, 0.14) |
| % death on the floor | 9.6e-07 (8.6e-07, 1.1e-06) | 0.019 (0.012, 0.025) | 0.16 (0.15, 0.18) |
| % death in the ICU | 1.5e-07 (4.6e-08, 2.5e-07) | 0.11 (0.079, 0.13) | 0.29 (0.25, 0.33) |
| % step up from floor to ICU | 0.46 (0.46, 0.46) | 0.72 (0.68, 0.76) | 0.59 (0.54, 0.63) |
| % step down to the floor | 0.014 (0.014, 0.014) | 0.13 (0.11, 0.15) | 0.16 (0.14, 0.18) |
| Triage time in ED (days) | 0.51 (-0.19, 1.2) | 0.2 (0.19, 0.22) | 0.25 (0.24, 0.26) |
| Average LOS on floor (days) | 2.2 (2.2, 2.2) | 7.5 (7.1, 8) | 10 (9.9, 11) |
| Average LOS in ICU (days) | 0.13 (0.13, 0.13) | 12 (11, 14) | 9.7 (8.7, 11) |

Table 2: Probabilities of transition and lengths of stay assuming exponential distributions on time to each competing risk.

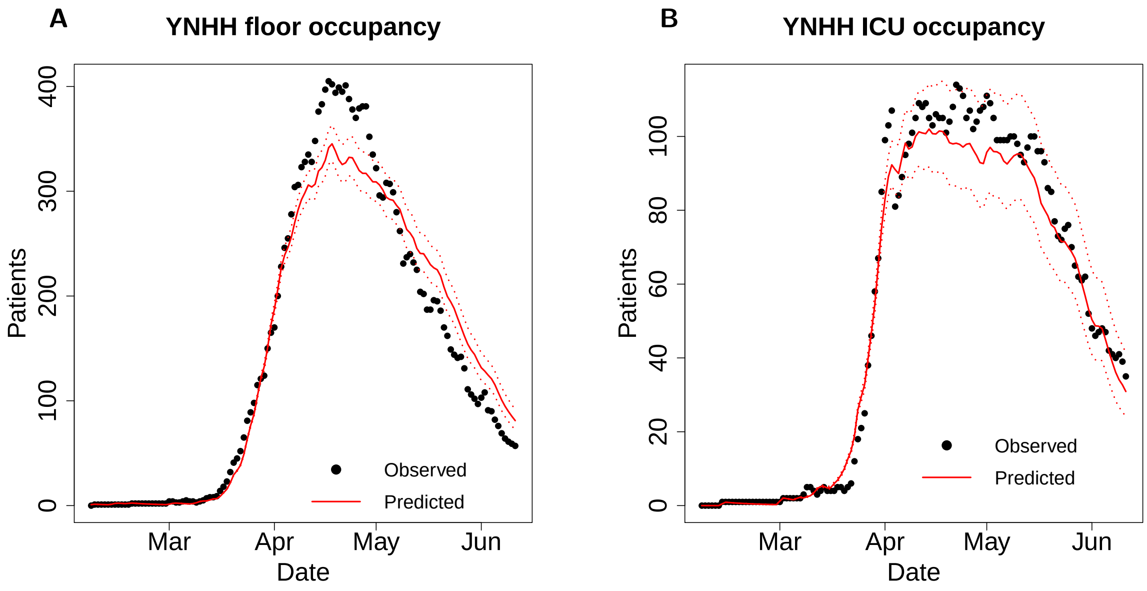


Figure 1: Inpatient predicted and observed COVID19 floor occupancy (A) and ICU occupancy (B). Parameters describing rates of transition between hospital departments were estimated assuming exponentially distributed time to event. The dotted line represents occupancy at YNHH. The solid red line represents occupancy predicted by the model based on parameters calculated using our fitting procedure and capacity estimates from YNHH. The dotted red lines represent estimated occupancy according to the bounds of 95% confidence intervals for each parameter.

#### 2. Gamma distribution for time to discharge from ED, floor, or ICU; exponential otherwise:

| | 0-18 years | 18-64 years | 65+ years |
|---|---|---|---|
| $\phi$ | 0.088 (0.088, 0.088) | 0.094 (0.094, 0.094) | 0.095 (0.095, 0.095) |
| $\sigma_{MS}$ | 1.1 (0, 5) | 0.37 (0, 2.3) | 0.55 (0.42, 0.68) |
| $\sigma_C$ | 0.072 (0, 0.32) | 4.1 (1.7, 6.6) | 0.48 (0.42, 0.55) |
| $\sigma_F$ | 0.48 (0, 1.7) | 4 (1.4, 6.6) | 3.1 (2.9, 3.2) |
| $\chi_C$ | 0.13 (0.13, 0.13) | 0.063 (0.055, 0.071) | 0.066 (0.057, 0.075) |
| $\chi_F$ | 0.027 (0, 0.098) | 0.11 (0.11, 0.12) | 0.071 (0.068, 0.075) |
| $\chi_{WF}$ | 0.027 (0, 0.098) | 0.11 (0.11, 0.12) | 0.071 (0.068, 0.075) |
| $\theta_F$ | 0.52 (0.52, 0.53) | 0.017 (0.014, 0.02) | 0.015 (0.014, 0.017) |
| $\theta_{WF}$ | 0.52 (0.52, 0.53) | 0.017 (0.014, 0.02) | 0.015 (0.014, 0.017) |
| $\eta$ | 36 | 4 36 | 36 |
| $\zeta$ | 5 (4.7, 5.6) | 1.2 (1.1, 1.3) | 0.92 (0.86, 0.99) |
| $\xi_{MS}$ | 0.012 (0.012, 0.012) | 0.0054 (0.0054, 0.0054) | 0.0039 (0.0039, 0.0039) |
| $\mu_{MS}$ | 0.00066 (0.00066, 0.00066) | 0.00066 (0.00066, 0.00066) | 0.00066 (0.00066, 0.00066) |
| $\mu_C$ | 6.3e-07 (6.1e-07, 6.6e-07) | 0.0083 (0.0061, 0.011) | 0.03 (0.025, 0.035) |
| $\mu_F$ | 7.6e-08 (0, 3.4e-06) | 0.0024 (0.0015, 0.0033) | 0.016 (0.014, 0.018) |
| $\mu_{WC}$ | 4 | 4 | 4 |
| $\mu_{WF}$ | 7.6e-08 (0, 3.4e-06) | 0.0024 (0.0015, 0.0033) | 0.016 (0.014, 0.018) |

Table 3: Parameters assuming exponential distributions for each competing risk, except for gamma distributed time to discharge.

| | 0-18 yrs | 19-64 yrs | 65+ yrs |
|---|---|---|---|
| Age distribution in ED | 0.02 (0.02, 0.02) | 0.58 (0.58, 0.58) | 0.4 (0.4, 0.4) |
| % discharged from ED | 0.69 (0.56, 0.81) | 0.17 (-0.26, 0.59) | 0.13 (0.11, 0.16) |
| % admitted from ED to floor | 0.27 (0.16, 0.39) | 0.56 (0.41, 0.71) | 0.75 (0.72, 0.78) |
| % admitted from ED to ICU | 0.037 (-0.0043, 0.086) | 0.27 (-0.00055, 0.54) | 0.12 (0.1, 0.13) |
| % death on the floor | 9.4e-07 (-2.5e-07, 2.1e-06) | 0.018 (0.012, 0.024) | 0.15 (0.14, 0.17) |
| % death in the ICU | 5.4e-07 (-1.2e-07, 1.2e-06) | 0.12 (0.089, 0.15) | 0.31 (0.27, 0.35) |
| % step up from floor to ICU | 0.28 (0.28, 0.28) | 0.8 (0.77, 0.83) | 0.62 (0.58, 0.66) |
| % step down to the floor | 0.31 (0.3, 0.31) | 0.13 (0.11, 0.14) | 0.15 (0.13, 0.17) |
| Triage time in ED (days) | 0.59 (-0.22, 1.4) | 0.16 (0.074, 0.24) | 0.24 (0.23, 0.26) |
| Average LOS on floor (days) | 0.35 (0.34, 0.36) | 7.3 (6.9, 7.8) | 9.6 (9.2, 10) |
| Average LOS in ICU (days) | 23 (23, 23) | 14 (12, 15) | 10 (9.2, 11) |

Table 4: Probabilities of transition and lengths of stay assuming exponential distributions on time to each competing risk, except for gamma-distributed time to discharge.

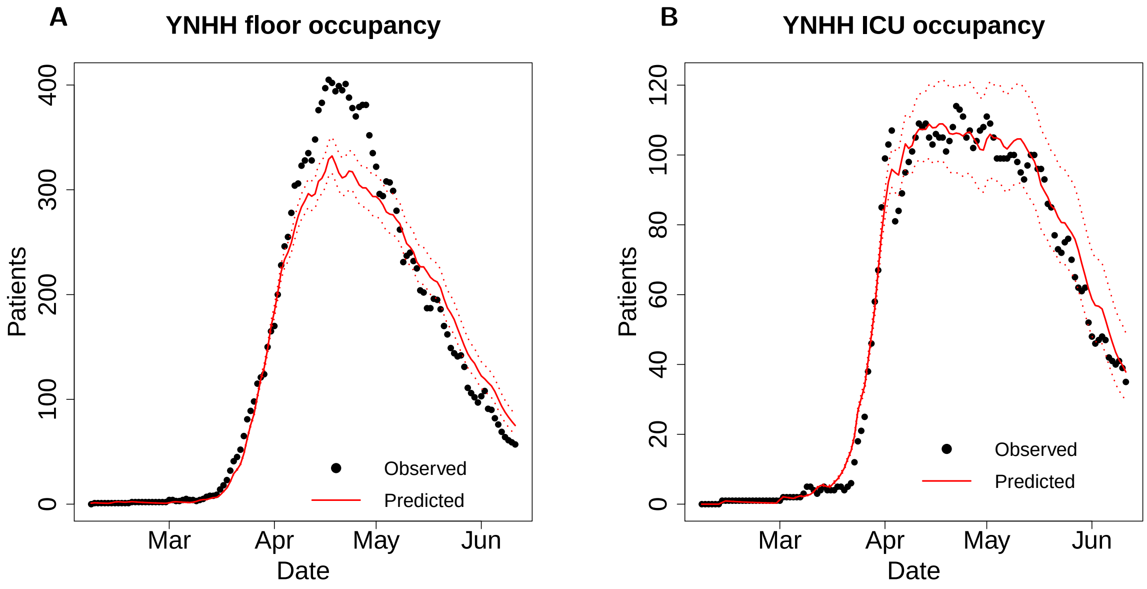


Figure 2: Inpatient predicted and observed COVID19 floor occupancy (A) and ICU occupancy (B). Parameters describing rates of transition between hospital departments were estimated assuming exponentially-distributed time to event, except for time to discharge in the ICU and on the floor, which were assumed to be gamma-distributed. The dotted line represents either occupancy or cumulative observed at YNHH. The solid red line shows occupancy predicted by the model based on parameters calculated using our fitting procedure and capacity estimates from YNHH. The dotted red lines represent estimated occupancy according to the bounds of 95% confidence intervals for each parameter.

### 3 Design and construction of the R shiny web application

A version of the model described in this report was implemented in the R shiny web application, deployed at `https://forrestcrawford.shinyapps.io/covid19_icu/`. Figure 3 shows the default page of the web application. This interactive web application was intended to be a tool for scenario analysis, used by hospital administrators and departments involved in capacity planning during the COVID-19 pandemic. This tool takes as an input an infection scenario which can be specified by the user, the capacity and current occupancy of the hospital system, and parameters describing basic features of the patient population served by the hospital. The tool allows the user to specify a strategy for capacity expansion, observing the effect of adding beds to the system. The outputs of the tool include projections of expected occupancy, deaths, an estimate of the time at which the system would reach capacity, and an estimate of the number of extra beds which would be necessary to relieve the overflow.

Because exact time series of COVID-19 presentations were not always easily accessible, we created an opening dashboard on the “Scenario" tab which allows the user to specify an infection scenario (Fig. 4). On this dashboard, the user can specify a time horizon for projections, the shape of the infection curve, and parameters which control the shape of this curve. The infection curve represents the number of new COVID-19 presentations to the health system each day, rather than the number of new COVID-19 infections in the population. Thus, the inputs to this model could be based on model fits to observed COVID-19 ED presentations at a particular hospital. This feature allows users to test various scenarios without having access to exact time series of ED presentations. Furthermore, these scenarios can be studied without knowledge of highly uncertain dynamics of new infections in the population.

##### 3.0.1 ED presentation dynamics

The functions for ED presentations and capacity are intentially left unspecified and can be set by the user. The user can choose among the following options:

- **Exponential increase:** The number of ED presentations rises exponentially over the time-period in question from its initial value, with an exponent of $\frac{\ln(2)}{T_{\text{doubling}}}$, with $T_{\text{doubling}}$ being the user-specified doubling time. This is especially relevant early-on in the epidemic.
- **Linear increase:** The number of ED presentations rises linearly over the time-period in question from its initial to its final value. This is relevant when the epidemic has not peaked but is being kept somewhat in check by various interventions.
- **Saturating:** In this case, the number of ED presentations plateaus to its final value at the end of the time-horizon, rising as a logistic function centered in the middle of the time period under study. This is relevant when the epidemic is close to peaking.
- **Flat:** The number of ED presentations remains equal to day zero across the time-frame.

The user can also model the effect of exponential, linear, and saturating decrease in ED presentations using the tool, for example due to the implementation of non-pharmaceutical interventions.

On the “Capacity” and “Strategy” tabs, the user can specify the capacity of their healthcare system and a strategy for capacity expansion. Defaults in the web application are based on YNHH capacity. The “Capacity" tab includes inputs for the number of floor and ICU beds available at baseline, as well as the percentage occupied at time zero for the intended simulation (Fig. 5). The “Strategy" tab can be toggled on and off depending on whether a strategy for further surge is planned (Fig. 6). When on, the user can specify a target number of beds for the floor and ICU, as well as the time frame within which the capacity expansion is expected to occur.

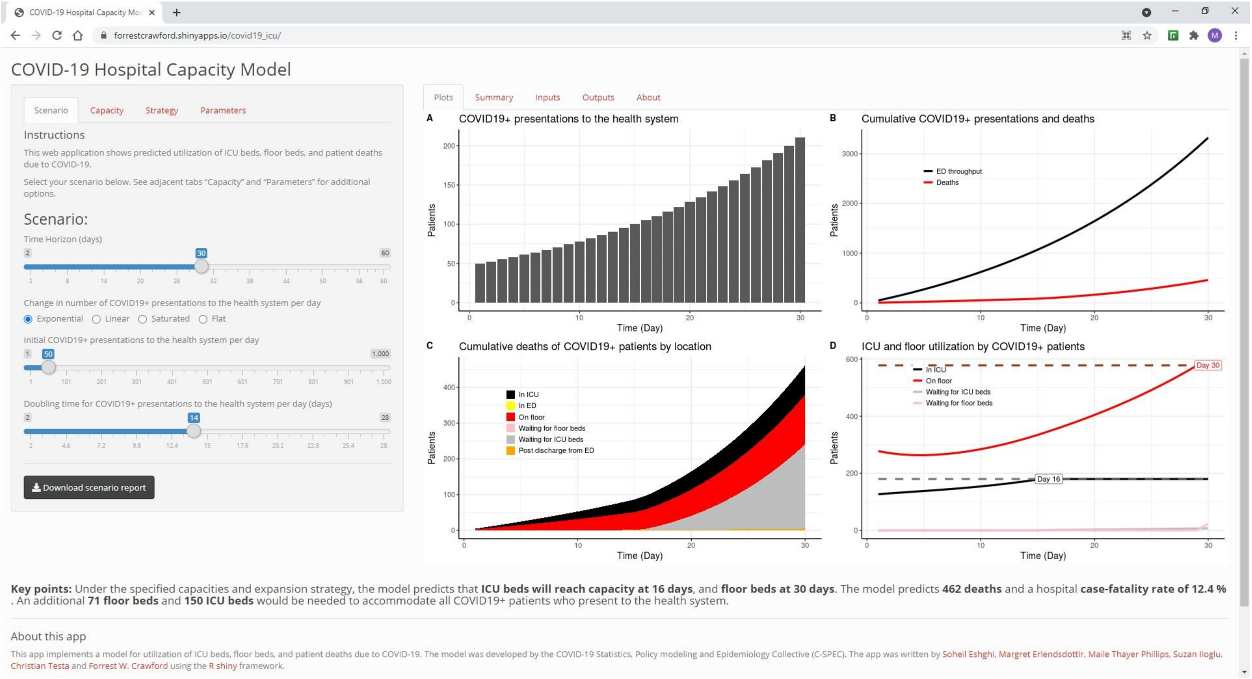


Figure 3: **Home page**: The default page of the web application, available at `https://forrestcrawford.shinyapps.io/covid19_icu/`

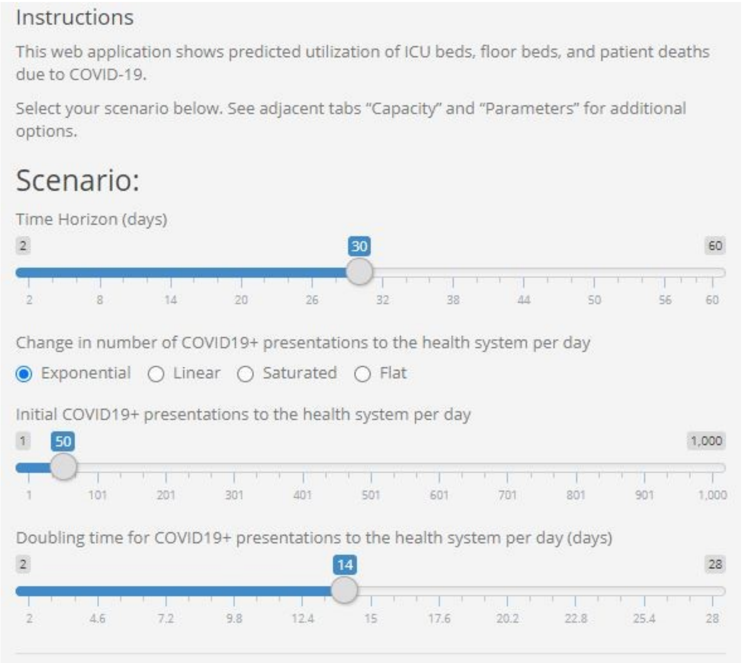


Figure 4: **Scenario tab:** allows the user to select a timeseries of daily COVID-19 presentations to a health system.

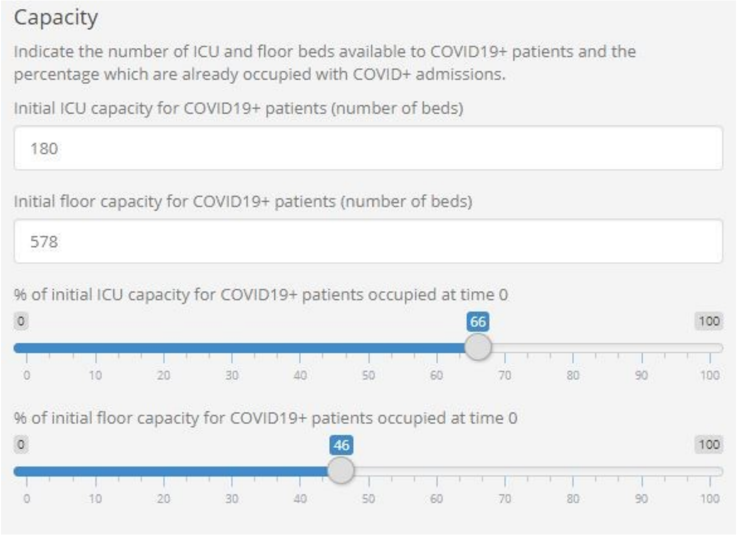


Figure 5: **Capacity tab:** allows the user to specify the number of beds available to COVID-19 patients, and the number of beds occupied at time zero.

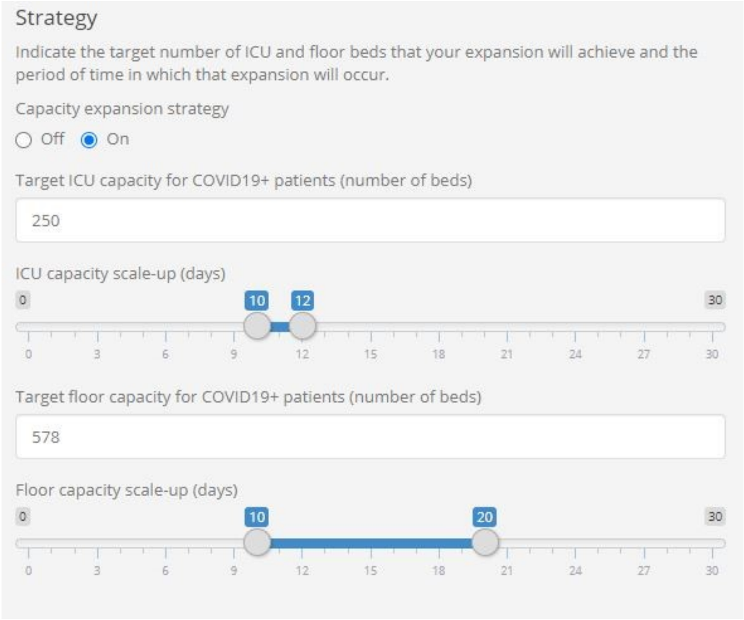


Figure 6: **Strategy tab:** allows the user to specify whether or not the health system plans to surge, when the surge would occur, and the number of beds planned.

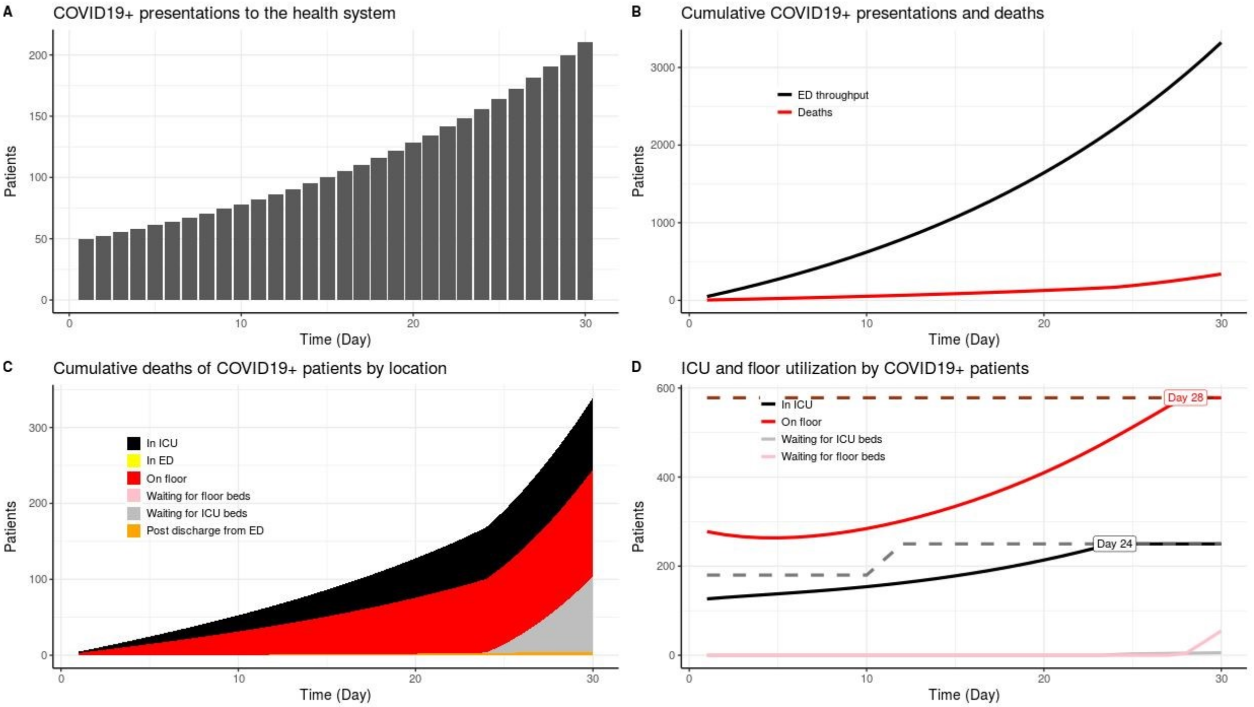


Figure 7: **Plots tab:** These plots are the main output of the model. Plot A shows the time series of presentations to the ED, Plot B shows cumulative ED presentations and projected cumulative deaths, and Plot C shows cumulative deaths by location. Plot D demonstrates projected occupancy on the floor and in the ICU, as well as a flag which appears with the time at which capacity of the healthcare system would be overwhelmed given the scenario and capacity settings entered by the user.

The “Parameters" tab allows the user to adjust several parameters which we determined to be easily estimated and important for tailoring model dynamics to specific patient populations. For example, overall admission and death rates are significantly different between age groups, and we have included a two-part slider which allows the user to control the age distribution of the population in the simulation. Furthermore, average LOS on the floor and in the ICU is tracked by hospitals and influences both occupancy and the time at which the hospital would reach capacity.

The model outputs are presented on the “Plots" and “Summary” tabs. We included “Key point” which include the most important outputs to hospital administrators, chosen based on feedback from YNHHS administrators. Of especial interest were the time to full capacity and the number of additional beds, if any, which would be necessary to accommodate all COVID-19 patients. On the “Plots” tab (Fig. 7), Plots A and B show daily and cumulative COVID-19 presentations to the health system. Plots B and C show cumulative deaths predicted by the model, as well as a breakdown of these deaths by department. Plot D shows the occupancy in the ICU and the floor predicted by the model, as well as flags which denote the time at which capacity is reached, if it is exceeded during the simulation. Exact values for anticipated deaths, days to ICU and floor overflow, extra beds needed, and predicted case-fatality rates are summarized on the “Summary” tab.

To aid new users in the use of the model, we have included three tabs - “Input”, “Outputs”, and “About” - which qualitatively describe the model inputs, outputs, and structure.

#### References


[1] Mattia Ferraresi. A coronavirus cautionary tale from Italy: Don’t do what we did. *The Boston Globe*, March 2020.

[2] Fei Zhou, Ting Yu, Ronghui Du, Guohui Fan, Ying Liu, Zhibo Liu, Jie Xiang, Yeming Wang, Bin Song, Xiaoying Gu, Lulu Guan, Yuan Wei, Hui Li, Xudong Wu, Jiuyang Xu, Shengjin Tu, Yi Zhang, Hua Chen, and Bin Cao. Clinical course and risk factors for mortality of adult inpatients with COVID-19 in Wuhan, China: a retrospective cohort study. *The Lancet*, 395(10229):1054–1062, March 2020.

[3] Preliminary Estimates of the Prevalence of Selected Underlying Health Conditions Among Patients with Coronavirus Disease 2019 - United States, February 12 - March 28, 2020. *Centers for Disease Control and Prevention, MMWR Morbidity Mortality Weekly Report*, April 2020.

[4] Severe Outcomes Among Patients with Coronavirus Disease 2019 (COVID-19) - United States, February 12 - March 16, 2020. *Centers for Disease Control and Prevention, MMWR Morbidity Mortality Weekly Report*, March 2020.

[5] Hospitalization Rates and Characteristics of Patients Hospitalized with Laboratory-Confirmed Coronavirus Disease 2019 - COVID-NET, 14 States, March 1-30, 2020. *Centers for Disease Control and Prevention, MMWR Morbidity Mortality Weekly Report*, April 2020.

[6] Characteristics and Clinical Outcomes of Adult Patients Hospitalized with COVID-19 - Georgia, March 2020. *Centers for Disease Control and Prevention, MMWR Morbidity Mortality Weekly Report*, May 2020.
